# Brain activity measured by functional brain imaging predicts breathlessness improvement during pulmonary rehabilitation

**DOI:** 10.1101/2021.11.26.21266908

**Authors:** Sarah L. Finnegan, Michael Browning, Eugene Duff, Catherine J. Harmer, Andrea Reinecke, Najib M. Rahman, Kyle T.S. Pattinson

## Abstract

**Background:** Chronic breathlessness in COPD is effectively treated with pulmonary rehabilitation. However, baseline patient characteristics predicting improvements in breathlessness are unknown. This knowledge may provide better understanding of the mechanisms engaged in treating breathlessness, helping to individualise therapy. Increasing evidence supports the role of expectation (i.e. placebo and nocebo effects) in breathlessness perception. In this study, we tested functional brain imaging markers of breathlessness expectation as predictors of therapeutic response to pulmonary rehabilitation, and whether D-cycloserine, a brain-active drug known to influence expectation mechanisms, modulates any predictive model.

**Methods:** Data from 72 participants with mild-to-moderate COPD recruited to a randomised double-blind controlled experimental medicine study of D-cycloserine given during pulmonary rehabilitation was analysed (ID: NCT01985750). Baseline variables, including brain-activity, self-report questionnaires responses, clinical measures of respiratory function and drug allocation were used to train machine-learning models to predict the outcome, a minimally clinically relevant change in the dyspnoea-12 score.

**Findings:** Only models that included brain imaging markers of breathlessness-expectation successfully predicted improvements in dyspnoea-12 score (sensitivity 0.88, specificity 0.77). D-cycloserine was independently associated with breathlessness improvement. Models that included only questionnaires and clinical measures did not predict outcome (sensitivity 0.68, specificity 0.2).

**Interpretation:** Brain activity to breathlessness related cues is a strong predictor of clinical improvement in breathlessness over pulmonary rehabilitation. This implies that expectation is key in breathlessness perception. Manipulation of the brain’s expectation pathways (either pharmacological or non-pharmacological) merits further testing in the treatment of chronic breathlessness.

**Funding:** This work was supported by the JABBS Foundation

**Research in context:** *Evidence before the study:* Despite considerable research we still do not know which patient characteristics predict clinical improvements in breathlessness following pulmonary rehabilitation. Recent evidence suggests that the brain processes associated with breathlessness-expectation play an important contributory role in breathlessness severity. However, this has never been examined as a predictor of pulmonary rehabilitation outcome. The ability to predict outcomes has a number of potential benefits, including identifying targets for personalised medicine and the better allocation of scare healthcare resources via parallel care pathways.

*Added value of the study:* This study analysed data from a longitudinal experimental medicine study of 71 patients with COPD over a course of pulmonary rehabilitation, that used functional magnetic resonance imaging testing breathlessness-expectation mechanisms in the brain. Participants were randomised in a double-blind procedure to receive either 250mg oral D-cycloserine or a matched placebo. Using baseline variables to train machine learning models we revealed that only models containing brain markers of breathlessness-expectation successfully predicted improvements in dyspnoea-12 score (sensitivity 0.88, specificity 0.77). D-cycloserine use was independently associated with breathlessness improvements. Models that only contained questionnaire and clinical measure did not predict outcome (sensitivity 0.68, specificity 0.2).

*Implications of all the available evidence:* These findings are the first evidence that breathlessness-expectation related brain activity is a strong predictor of clinical improvement in breathlessness over pulmonary rehabilitation. This implies that expectation is a key mechanism in breathlessness perception and that the manipulation of the brain’s expectation pathways merits further testing as a novel therapeutic approach for breathlessness.

## Introduction

Chronic breathlessness is a key feature of chronic obstructive pulmonary disease (COPD) with symptoms often persisting despite maximal medical therapy. Pulmonary rehabilitation is the best treatment for chronic breathlessness in COPD [1] but the response is variable. Thirty percent of people who complete pulmonary rehabilitation derive no clinical benefit [2]. Despite considerable research, we still do not know which patient characteristics predict beneficial response to pulmonary rehabilitation [2-5]. The ability to predict outcome has a number of potential benefits. These include improving our understanding of underlying mechanisms, identifying targets for personalised medicine, and may allow more accurate allocation of scarce healthcare resources.

Breathlessness severity is often poorly explained by objective clinical measures [6]. This has prompted research into identifying brain perceptual mechanisms that may explain this discordance. A body of work has recently identified that brain processes relating to expectation (akin to placebo and nocebo effects) have an important role in contributing to breathlessness severity. Whether brain-derived metrics may help predict outcome from pulmonary rehabilitation is unknown, and prediction models until now have not included measures of expectation.

Between-subject variability in therapeutic response is increasingly recognised as a confounder in clinical trials. A personalised medicine approach aims to identify subgroups of patients that respond to a specific therapy. In psychiatry, brain-derived metrics using functional neuroimaging have taken similar approaches to identifying subtypes of depression that may respond to bespoke therapies [7]. Such techniques rely on biomarkers, which may consist of predictive combinations of biochemical, genetic, demographic, physiological or cognitive measures. In the context of treating breathlessness, predictive biomarkers could pave the way for novel pharmacological and non-pharmacological treatments. These may either work as standalone therapies, or by enhancing other therapies, such as pulmonary rehabilitation.

In this study, we aimed to predict improvements in breathlessness during pulmonary rehabilitation by analysing baseline data from a longitudinal experimental medicine study of D-cycloserine upon breathlessness during pulmonary rehabilitation. We selected D-cycloserine, which is a partial agonist at the NMDA receptor in the brain, for its action on neural plasticity and influence on brain expectation mechanisms associated with cognitive behavioural therapies [8-10]. Brain based pharmacological adjuncts may be one opportunity to boost the effects of pulmonary rehabilitation. We hypothesised baseline brain activity in response to breathlessness-related expectation would predict improvement in breathlessness over pulmonary rehabilitation, and that if D-cycloserine indeed had an effect upon expectation then it would emerge as a significant factor in the prediction model. Given that moderators of treatment success of pharmacological agents such as D-cycloserine remain unclear, this information will not only help us build a better picture of the brain-behaviour changes that may underly response to pulmonary rehabilitation and therefore its potential value as a therapeutic target.

## Methods and Materials

A brief overview of materials and methods is presented here with full details included within the supplementary materials. The study and statistical analysis plan were pre-registered on bioXIV (https://osf.io/bfqds/). This was an analysis of data from a longitudinal experimental medicine study of patients with COPD over a course of pulmonary rehabilitation. Parts of the study were first published in a characterisation of baseline patient clusters [11] and subsequently in the investigation of the effect of D-cycloserine on brain activity (preprint - https://tinyurl.com/3pyupwkj). The analysis conducted for this study is novel, not previously reported and is the first use of predictive analysis using this dataset.

### Participants

71 participants (18 female, median age 71 years (46-85 years)) (Supplementary Table 1) were recruited immediately prior to enrolment in a National Health Service-prescribed course of pulmonary rehabilitation. Written informed consent was obtained from all participants prior to the start of the study. Study approval was granted by South Central Oxford REC B (Ref: 118784, Ethics number: 12/SC/0713). Full demographic details are included within the supplementary materials and are published separately (preprint is available at https://tinyurl.com/3pyupwkj).

### Study Protocol

Data for this analysis were acquired at baseline assessment held at the start of a pulmonary rehabilitation course, and following completion of the pulmonary rehabilitation at 6-8 weeks. At each study visit, identical measures were collected. Following the first visit, participants were randomized in a double-blind procedure to receive either 250mg oral D-cycloserine or a matched placebo. Participants received a single dose on four occasions 30 minutes prior to the onset of the first four pulmonary rehabilitation sessions.

#### Self-report questionnaires

All questionnaires were scored according to respective manuals: Dyspnoea-12 (D12) Questionnaire [12], Centre for Epidemiologic Studies Depression Scale (CES-D) [13], Trait Anxiety Inventory (TRAIT) [14], Fatigue Severity Scale [15], St George’s Respiratory Questionnaire (SGRQ) [16], Medical Research Council (MRC) breathlessness scale [17], Breathlessness catastrophising scale, adapted from the Catastrophic Thinking Scale in Asthma [18], Breathlessness vigilance, adapted from the Pain Awareness and Vigilance Scale Breathlessness Awareness and Vigilance Scale [19].

#### Physiological Measures

Spirometry and two Modified Shuttle Walk Tests (MSWT) were collected using standard protocols [20, 21]. Participant height and weight were recorded at each visit. Arterial oxygen saturations were collected at rest and following the MSWT.

### MRI Measures

#### Image acquisition

Magnetic resonance imaging of the brain was carried out using a Siemens 3T MAGNETOM Trio. A T1-weighted (MPRAGE) structural scan (voxel size: 1 × 1 × 1 mm) was collected and used for registration purposes. A T2*-weighted, gradient echo planar image (EPI) scan sequence (TR, 3000ms; TE 30ms; voxel size: 3 × 3 × 3 mm) was used to collect functional imaging data during the word cue task.

#### Word cue task

Given sufficient fearful breathlessness exposures, the suggestion alone of the situation can be sufficient to drive a top-down neural cascade and produce breathlessness in the absence of afferent inputs. We drew on this link to probe the neural responses of breathlessness-related expectation by examining the activity of brain regions responding to breathlessness-related word cues [22, 23]. Brain activity was correlated with corresponding visual analogue ratings of breathlessness and breathlessness-anxiety collected during scanning. During the fMRI scanning, participants were presented with a word cue, e.g., “climbing stairs” in white text on a black background for 7 seconds. Participants were then asked, “how breathless would this make you feel” (wB) and “how anxious would this make you feel” (wA). To each question participants responded within a 7 second window using a button box and visual analogue scale (VAS). The response marker always initially appeared at the centre of the scale, with the anchors “Not at all” and “Very much” at either end. Scan duration was 7 minutes and 33 seconds.

## Analysis

### Regions of interest

15 regions of interest were selected a-priori (Figure 1), encompassing regions associated with sensory and affective processing of breathlessness as well as body, symptom perception and emotional salience [22, 24, 25]. Regions were defined by standard anatomical atlas maps (Harvard-Oxford Atlas and Destrieux’ cortical atlas), thresholded at 40% probability and binarized.

**Figure 1.**
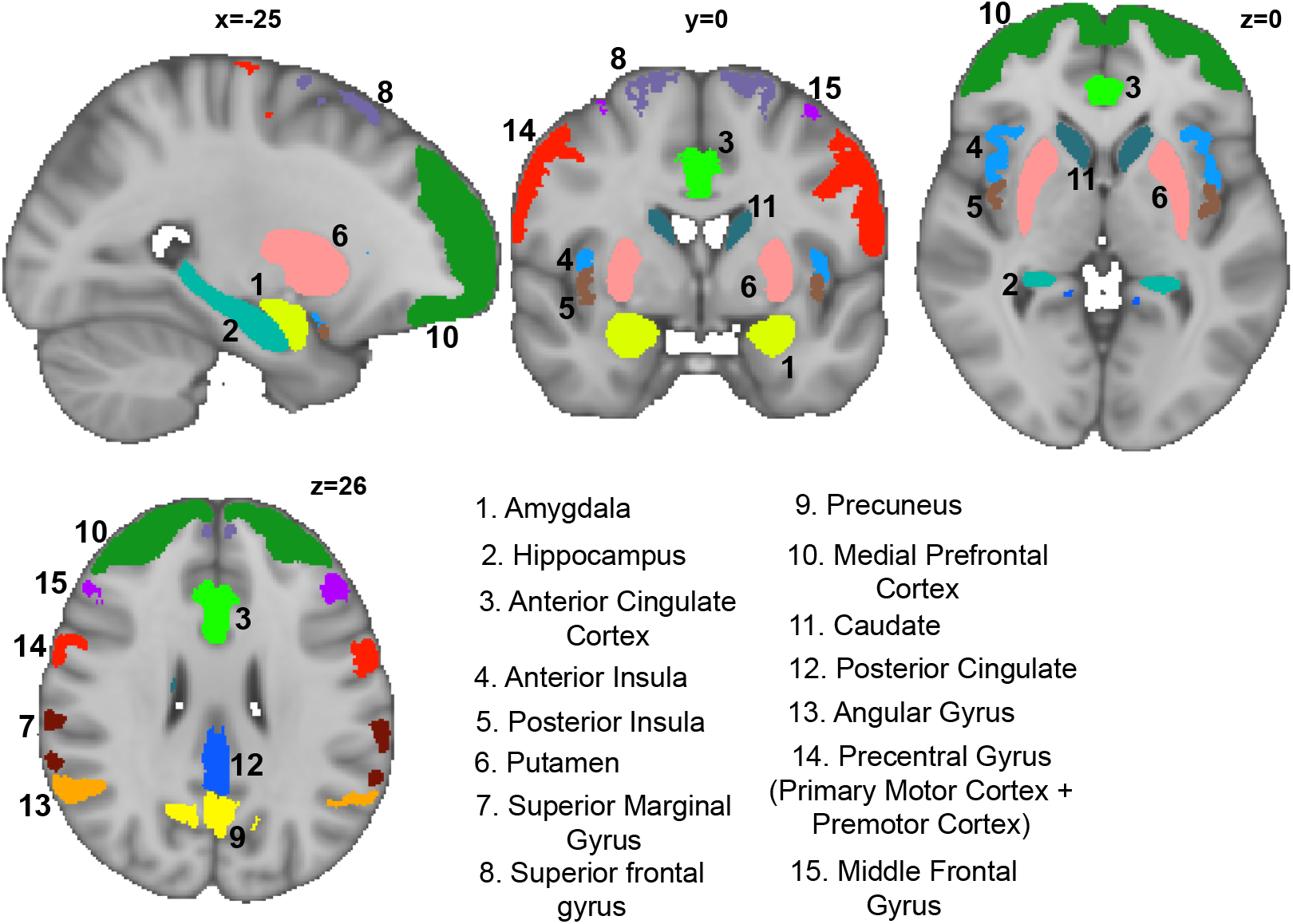
Region of interest map showing 15 brain areas.

#### Brain Imaging Analysis

Image processing was carried out using the Oxford Centre for Functional Magnetic Resonance Imaging of Brain Software Library (FMRIB, Oxford, UK; FSL version 5.0.8; https://ww.fmrib.ox.ac.uk/fsl/), MATLAB R2018b (Mathworks, Natick, MA) and associated custom scripts. Functional MRI processing was performed using FEAT (FMRI Expert Analysis Tool, within the FSL package).

#### Pre-processing and single subject models

Data were pre-processed according to standard protocols which included motion correction and physiological noise removal, before being entered into single subject general linear models. These models captured brain activity during the periods in which the breathlessness-related word cues were presented allowing us to examine expectation-related processes (Supplementary Figure 3). Further details regarding pre-processing and model specifics can be found within the supplementary materials. For each participant the mean signal in response to the breathlessness-related word cues was extracted for each brain region (Figure 1). This gave each participant 15 brain-derived scores to enter into the predictive models.

### Definition of response to pulmonary rehabilitation

Responsiveness to pulmonary rehabilitation was defined as a change in D12 score, a well-validated clinical measure of breathlessness, of three or more points, consistent with the minimal clinically important difference (MCID) [12]. To examine whether baseline D12 score differed significantly between responders and non-responders to pulmonary rehabilitation we conducted a comparison of mean baseline D12 score. In addition, a single logistic regression model was applied to the data using MATLAB’s mnrfit function to examine whether baseline D12 explained pulmonary rehabilitation outcome over and above the best model prediction. Significance was set as False Discovery Rate (FDR) corrected p<0.05.

### Predictive Models

#### Physiological Measures

Spirometry and two Modified Shuttle Walk Tests (MSWT) were collected using standard protocols [20, 21]. Participant height and weight were recorded at each visit. Sex was self-reported. Arterial oxygen saturations were collected at rest and following the MSWT.

Models were programmed using R version 3.6.1 (2019-07-05). Modelling procedure remained the same for each of the three models (Figure 2).

1. All measures were centred and scaled. Checks were performed to determine whether any measures were highly correlated (R>0.8) or linear combinations of each other (Supplementary Figure 3).
2. To correct for imbalance in the number of responders/non-responder a resampling procedure. Imbalanced classes can affect classifier performance. Random OverSample Examples (ROSE) was carried out. ROSE, an R package, creates an artificially balanced sample using a smoothed bootstrap approach [26].
3. An elastic net procedure was used to identify the number most relevant features for inclusion into the model. Elastic net procedure was selected for its ability to regularise, improve data sparsity via feature selection and cluster correlated measures together (for more details see supplementary materials). Features were selected based on ranked coefficients.
4. Model training parameters – C, kappa and sigma were selected based on an internal repeated cross-validation procedure (10-fold cross validation repeated 3 times). In all instances automated tuning parameter selection for the values, with a tune length of 5, was utilised within R’s *caret* package. Train-test data were kept separate across folds, with the algorithm never having access to the entire dataset. The best tuning parameters were selected automatically by R’s *caret* package from across cross validation folds.
5. These parameters were used to train a Support Vector Classifier (SVC) with radial kernel to predict outcomes in the entire dataset.
6. Model performance was assessed internally using accuracy, sensitivity, specificity and area under the curve (AUC). Full confusion matrices are presented. Model significance was assessed with a one-tailed binomial test of model accuracy compared to the null information rate.

**Figure 2.**
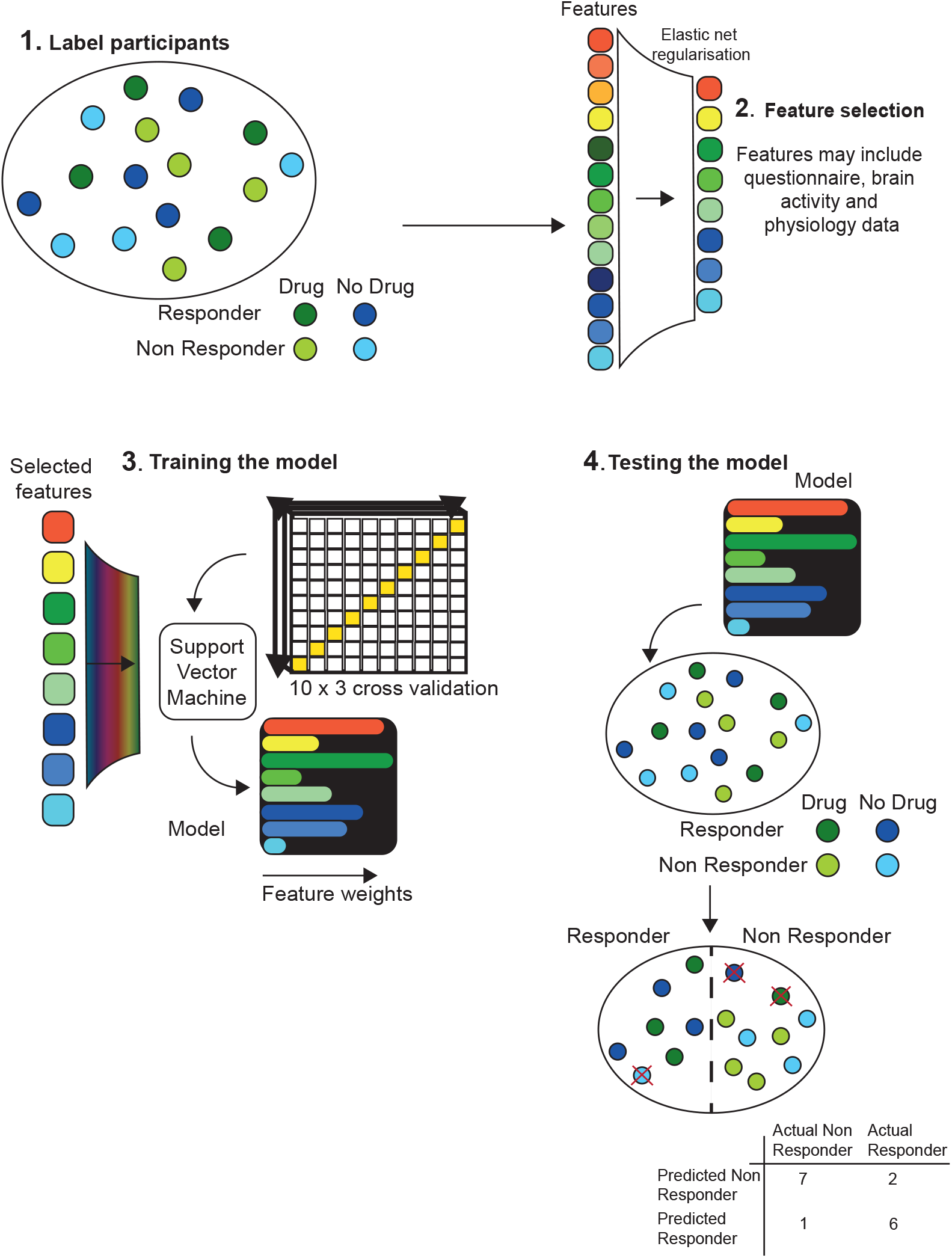
A schematic of the modelling procedure adapted from an illustration by Chekroud and colleagues [27]. **1**. Participants received two labels, the first corresponding to drug or placebo and the second “responder” or “non-responder” to treatment. **2**. An elastic net procedure was used to rank and select the top features. **3**. Selected features were used to develop model training parameters in a repeated-cross validation procedure in which the algorithm never has access to the entire dataset. **4**. The Trained SVM was then provided with the entire dataset to classify. In addition to model statistics, full confusion matrices were output to assess sensitivity and specificity.

## Results

### Responders and non-responders

41 / 71 participants in the primary dataset met the criteria of a change in D12 score of three or more points to be considered a responder [12] (24 D-cycloserine, 17 placebo), and 30 did not (13 D-cycloserine, 17 placebo). No significant interaction between responders and non-responders and drug was identified using Chi squared analysis ((1,N=71) = 1.6, p=0.21).

### Feature selection – brain imaging only model

The elastic net procedure identified 13 of 15 brain derived metrics and drug as relevant for model inclusion. These features were: brain activity within amygdala, caudate, prefrontal cortex, hippocampus, superior frontal gyrus, anterior insula, drug, posterior cingulate cortex, putamen, posterior insula, middle frontal gyrus, precuneus, precentral gyrus and angular gyrus.

### Feature selection – brain and non-imaging measure model

The elastic net procedure identified 12 of 15 brain derived metrics, 13 of 20 non-imaging measures including drug as relevant for model inclusion. These features were brain activity within: superior frontal gyrus, hippocampus, angular gyrus, superior marginal gyrus, amygdala, prefrontal cortex, precuneus, anterior cingulate cortex, anterior insula, middle frontal gyrus, posterior insula and putamen. Behavioural features identified as relevant for model inclusion were D12, anxiety, depression, MRC, the three St George’s domains (active, impact, symptoms), MWST BORG, heart rate and Sats change, fatigue, age and BMI.

### Feature selection – non-imaging measures model

Of the 20 questionnaire and physiological features available, only D12 survived the feature selection process.

### Model results – Internal validation

Three models with variables selected by the elastic net procedure were assessed for their ability to discriminate responders from non-responders (Table 3).

**Table 1.**
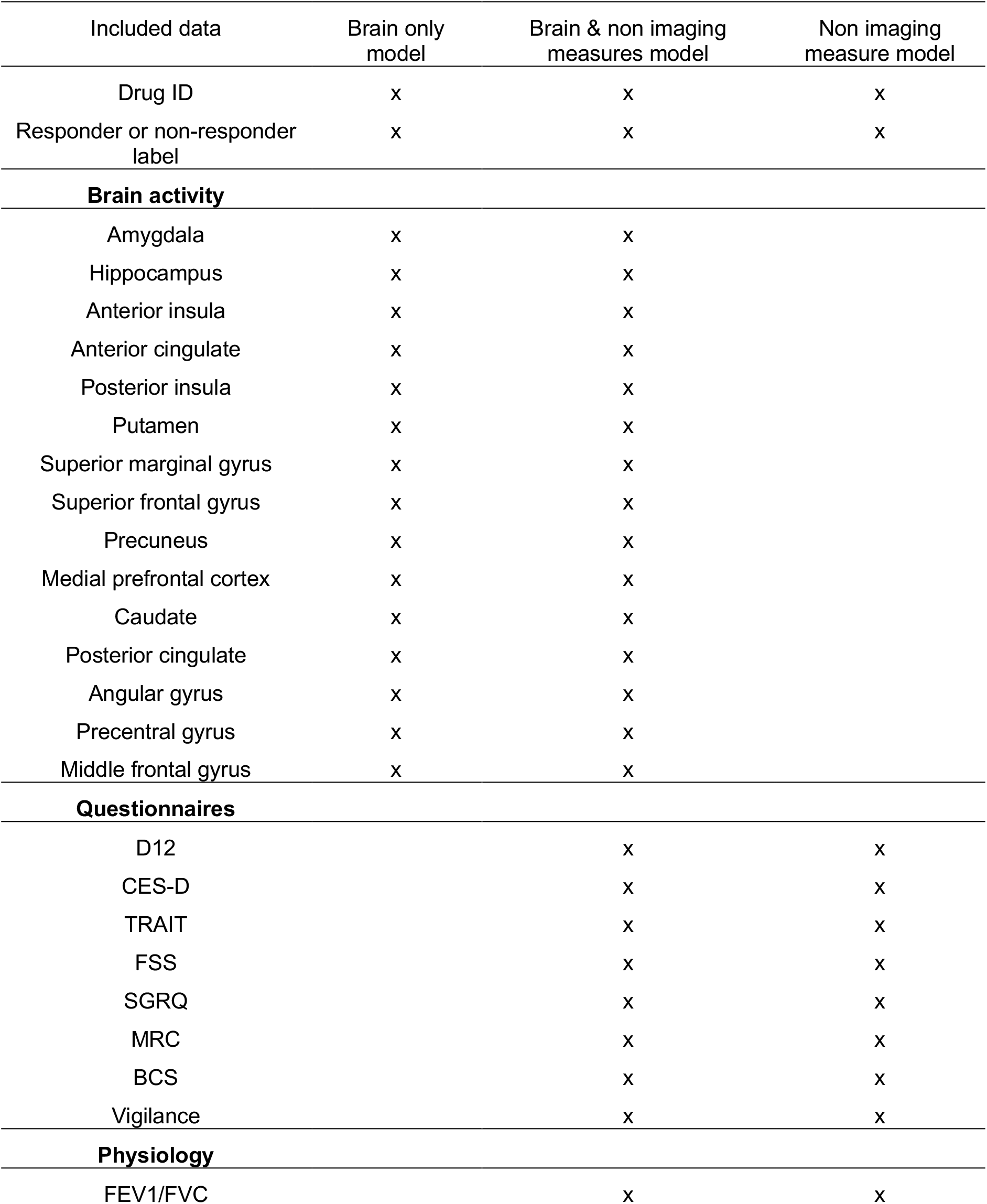
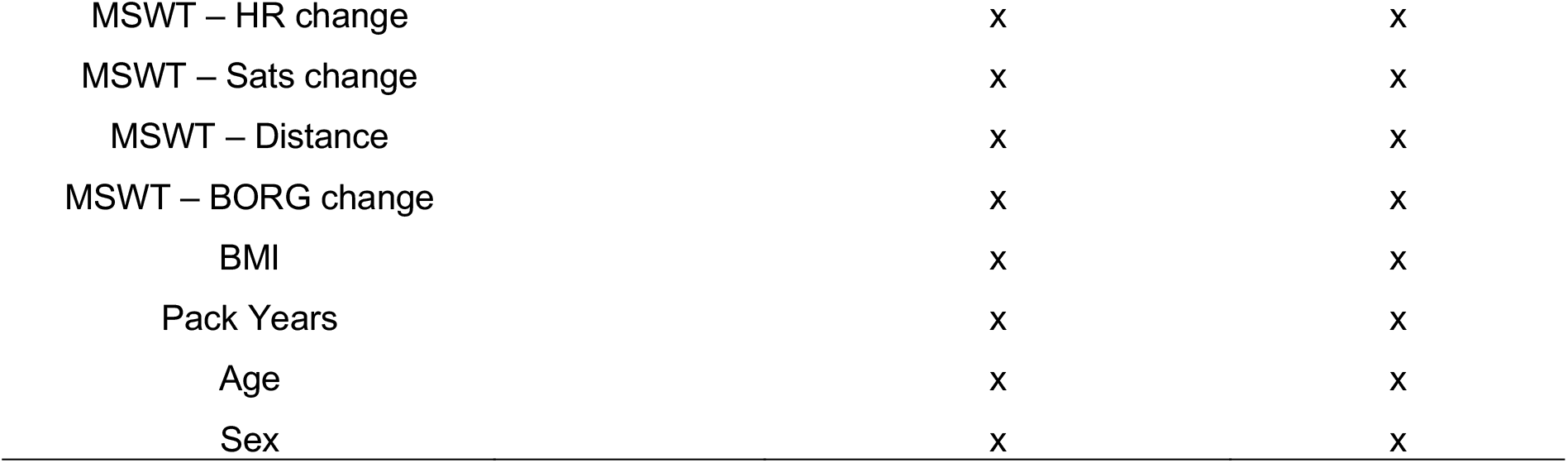
List of measures included within each of the three models (indicated by “x”). Drug ID labels corresponded to whether the participant received D-cycloserine or placebo. Dyspnoea-12 (D12), Centre for Epidemiologic Studies Depression Scale (CES-D), Trait Anxiety Inventory (TRAIT), Fatigue Severity Scale (FSS), St George’s Respiratory Questionnaire (SGRQ), Medical Research Council (MRC) breathlessness scale, Breathlessness catastrophising scale (BCS), Heart Rate (HR), Oxygen Saturation (Sats).

**Table 2.** Logistic regression coefficients for predictive power of the computationally derived brain-behaviour model (model prediction labels), and baseline D12 on pulmonary rehabilitation outcome, measured as a change in D12 score above the minimal clinical important difference. Significance is expressed as False Discovery Rate (FDR) -corrected p-values.

|  | Coefficient | P value |
| --- | --- | --- |
| Intercept | 3.571 | 0.007 |
| Model prediction labels | -2.809 | <0.001 |
| Baseline D12 | 0.071 | 0.191 |

**Table 3.** Model statistics for brain imaging only, brain and non-imaging measure models and non-imaging measures only model. All models contained Drug ID as an additional term. AUC – area under the curve. CI – confidence interval. P-value is expressed as the result of a one-tailed binomial test of model accuracy compared to the null information rate.

|  |  | Brain only full model |  | Brain & non-imaging measures full model |  | Non-imaging measures full model |  |
| --- | --- | --- | --- | --- | --- | --- | --- |
| Accuracy |  | 0.70 |  | 0.83 |  | 0.66 |  |
| 95% CI |  | 58-81 |  | 0.72-0.90 |  | 0.54-0.77 |  |
| Sensitivity |  | 0.93 |  | 0.88 |  | 0.68 |  |
| Specificity |  | 0.40 |  | 0.77 |  | 0.20 |  |
| AUC |  | 0.79 |  | 0.87 |  | 0.70 |  |
| p-value |  | 0.02* |  | <0.001** |  | 0.09 |  |
| Confusion matrices |  | Reference |  | Reference |  | Reference |  |
|  |  | Yes | No | Yes | No | Yes | No |
| Prediction | Yes | 38 | 18 | 36 | 7 | 28 | 11 |
|  | No | 3 | 12 | 5 | 23 | 13 | 19 |

The combination of brain and behaviour metrics produced the best classification performance (accuracy - 0.83 (95% CI 0.75-0.90); sensitivity – 0.88; specificity – 0.77; p<0.001) (Table 3). Weighted variable importance was found to be similar across features, as demonstrated by the thickness of the lines in Figure 3. The brain only model was able to correctly categorise participants with statistically significant likelihood (accuracy 0.70 (95% CI 0.58-0.81).

**Figure 3.**
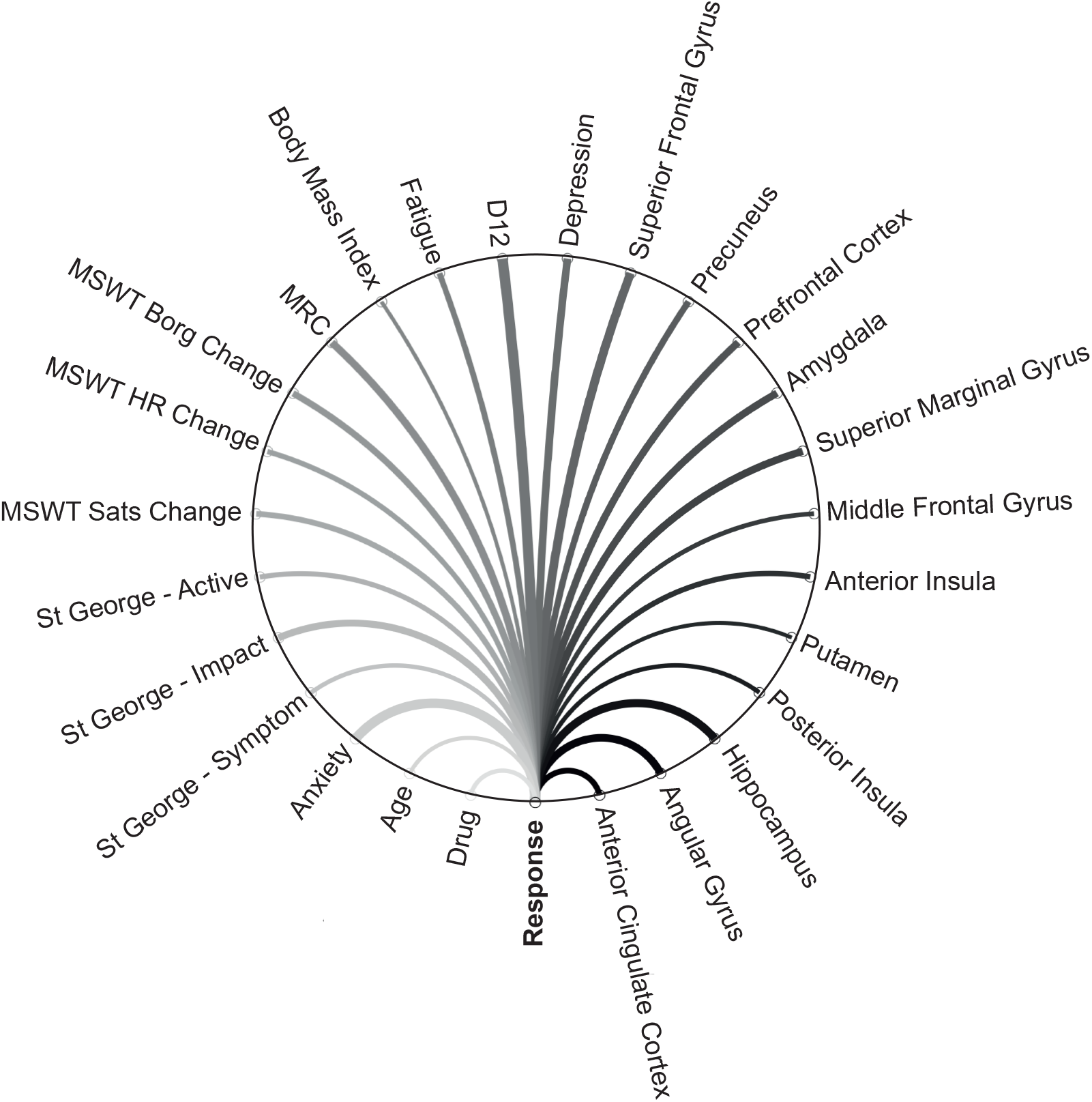
Schematic representation of the best predictive model. Predictive brain imaging and non-imaging measures are shown linked to treatment response by weighted lines, indicating variable importance. **MSWT –** Modified Shuttle Walk Test; **Sats –** Oxygen saturation; **HR –** Heart Rate; **MRC –** Medical Research Council;

## Discussion

### Key Findings

Using supervised machine learning, this study successfully identified markers that predict clinically relevant improvements in breathlessness over a course of pulmonary rehabilitation. The best model combined brain-imaging markers of breathlessness-expectation, self-report questionnaires and physiology measures, and demonstrated high sensitivity and specificity. Whether or not a participant received D-cycloserine was a significant feature in this model. Our findings demonstrate the first predictive model of change in breathlessness across pulmonary rehabilitation and, for the first time, the clinical relevance of expectation-related brain activity as a therapeutic target in the treatment of breathlessness.

To date, no study has produced a model capable of predicting an individual’s change in breathlessness over pulmonary rehabilitation from baseline traits [2-4]. Although previous studies have shown correlations between baseline variables and outcomes [5], none have attempted to predict outcomes at an individual level. This study is therefore the first to directly predict an individual’s change in breathlessness over pulmonary rehabilitation. This was achieved using sensitive brain imaging techniques in order to capture personalised responses to breathlessness expectation which has, until recently remained relatively unexplored.

Expectation has been linked with symptom severity across conditions including breathlessness and pain [28, 29], and is well recognised to underly the placebo and nocebo effects. An example of the nocebo effect in breathlessness is provided by a study of healthy volunteers in which, using a conditioning paradigm, a harmless odour was initially paired with induced breathlessness. Subsequently, the odour alone was shown to drive brain activity in the periaqueductal gray and anterior cingulate cortex leading to breathlessness despite the absence of afferent respiratory input [29]. In Abdallah et al [30], expectation-related brain activity was associated with poorer responses to opioids in breathlessness, potentially explaining why clinical trials of opioids in the management of breathlessness have been unsuccessful [31, 32].

Fear and anxiety are key components of expectation, which recent research suggests may play a key role in the mechanisms and maintenance of breathlessness [22, 33, 34]. Despite this, expectation-related effects have not previously been considered in prediction studies of pulmonary rehabilitation outcome. Our previous work showed a clear correlation between expectation-related brain activity in areas that include the anterior insula, anterior cingulate cortex and prefrontal cortex, and improvements in breathlessness over pulmonary rehabilitation [22]. However, while these studies suggest baseline cognitive state may be a therapeutically relevant target, importantly, the methods employed so far did not attempt to predict the response of an individual. Taken together, converging lines of evidence point towards expectation-related processes as a clear potential therapeutic target.

In this study we focused on brain activity changes within a set of pre-selected regions of interest associated with breathlessness-expectation and body and symptom perception [22, 24, 25]. In the original trial we hypothesised that D-cycloserine would augment the therapeutic effects of pulmonary rehabilitation across this network, via its effects on neural plasticity and promotion of expectation related learning [10, 35].

Using data driven techniques, 13 of the 15 brain derived metrics (and drug) were identified as relevant for model inclusion. Selected brain areas spanned the components of relevant body and symptom perception and emotional salience networks. The resulting brain-only model, while statistically significant (p=0.02) and possessing good sensitivity (0.93), did not distinguish responders from non-responders with sufficient specificity (0.40).

By, enriching the brain-only models with questionnaire and physiology measures improved performance considerably. In this enriched model, 12 brain derived metrics and 13 non-imaging derived metrics, which included self-report questionnaire measures, physiology and drug, were identified as relevant for model inclusion. Measures of accuracy (0.83), sensitivity (0.88) and specificity (0.77) all suggest this model was able to significantly (p<0.001) predict pulmonary rehabilitation outcome.

Within the non-imaging measure only model, D12 alone was selected by the elastic net and was not found to be significantly (p=0.09, sensitivity=0.68, specificity=0.20) predictive of pulmonary rehabilitation outcome. No other of the 13 non-imaging derived metrics available was found to contribute to the model. That only D12 was selected suggests that the remaining measures, which were important predictors of rehabilitation outcome in the enriched model, interact strongly with brain activity. These results highlight the value of approaching breathlessness from a multimodal data perspective.

The retained brain activity features implicate a range of brain networks encompassing functions of cognitive control, symptom perception and sensory integration. Activity within these regions has been shown to predict outcome to cognitive behavioural therapy in social anxiety disorder [36] and obsessive-compulsive disorder [37]. In our paradigm, in which patients were shown breathlessness related word-cues, triggering expectation related processes, activity within cognitive control network may indicate the allocation of attentional resources. The retained questionnaires within the brain and non-imaging measure model together highlight another important domain of breathlessness: symptom perception. We suggest that baseline symptom perception may act to directly influence the interpretation of the new experiences of pulmonary rehabilitation as positive or negative.

D-cycloserine has shown promise in trials examining anxiety, posttraumatic stress disorder and other mental health conditions [8-10], where acute dosages administered prior to exposure based therapies appear to augment changes to expectation, boosting therapeutic effects as a result. As a drug which acts on expectation-related brain activity pathways it is therefore not surprising that whether a participant received D-cycloserine or placebo was a retained as a feature in both the brain only model, and to a lesser extent the brain and non-imaging model.

### Limitations and future work

The major limitation of this study is the lack validation of the model in an external dataset. While some studies hold out a proportion of the original data to create an external validation dataset, this technique was not possible here due to restrictions of sample size. To address these limitations, we used a cross validation approach in which the support vector machine was exposed to multiple iterations of the sub-sampled dataset during model training, and therefore never “saw” the entire dataset until the test phase. Models with a large number of measures compared to events (responder or non-responder) risk overfitting and demonstrate poor generalisability to novel datasets. Our dataset contained 35 potential features and therefore was at risk of overfitting. To address this issue, we reduced the number of data-dimensions via feature selection, employed cross-validation and used an automated tuning of the regularisation parameter “C”. However, while these techniques may ameliorate some of the risk of overfitting, a future study with larger sample size, or independently collected datasets, would take the next steps to externally validate the brain-behaviour model and allow assessment of generalisability. Finally, a key feature of support vector machines is that they fit high-dimensional discriminatory planes between multiple measures to predict an outcome. While this multivariate approach does afford greater sensitivity in distinguishing between non-separable distributions, it also leads to less intuitive interpretations. Assigning directions to the relationships between variables or onto outcomes is not possible with this technique. Thus, the researcher must plan to scrutinise models via interventional studies.

While larger sample sizes are now required to translate these mechanistic models into clinical relevance, the data provides evidence that breathlessness expectation related brain activity at baseline strongly influences how patients respond to treatment in a predictable manner. Further work is now needed in order to move towards model validation and eventual clinical application.

## Conclusions

This study offers the first steps towards brain-based predictive biomarkers for pulmonary rehabilitation outcome. We have shown that by models including objective brain markers of breathlessness-expectation are able to predict, for the first time, which patients will have clinically important improvements in breathlessness over pulmonary rehabilitation. Such models could reduce the burden of complex decisions placed on the clinicians and pave the way for targeted behavioural and pharmacological interventions.

## Data Availability

All data produced in the present study are available upon reasonable request to the authors

## Author contributions

**S.L.F –** Acquisition of data, analysis, interpretation, drafting, editing and approving manuscript

**M.B** – Approval of statistical methods, editing and approving manuscript

**E.D** – Approval of statistical methods, editing and approving manuscript

**C.J.H –** Data interpretation, editing and approving manuscript

**A.R –** Data interpretation, editing and approving manuscript

**N.J.R –** Study interpretation, editing and approving manuscript

**K.T.S.P -** Study design, interpretation, editing and approving manuscript

## Declaration of interests

Dr. Harmer has valueless shares in p1vital and serves on their advisory panel. She has received consultancy payments from p1vital, Zogenix, J&J, Pfizer, Servier, Eli-Lilly, Astra Zeneca, Lundbeck. Dr. Pattinson is named as co-inventors on a provisional U.K. patent application titled “Use of cerebral nitric oxide donors in the assessment of the extent of brain dysfunction following injury. Dr. Rahman, has received consulting fees from Rocket Medical U.K. Dr Browning has received travel expenses from Lundbeck for attending conferences, has shares in P1vtial Products Ltd and has acted as a consultant for Jansen and for CHDR. The remaining authors have no biomedical financial interests or potential conflicts of interest.

## Funding

This work was supported by the JABBS Foundation. This research was funded in whole, or in part, by the Wellcome Trust 203139/Z/16/Z. For the purpose of Open Access, the author has applied a CC-BY public copyright licence to any Author Accepted Manuscript version arising from this submission.

CJH is supported by the National Institute for Health Research Biomedical Research Centre based at Oxford Health NHS Foundation Trust and The University of Oxford, and by the UK Medical Research Council. KTSP and NMR are supported by the National Institute for Health Research Biomedical Research Centre based at Oxford University Hospitals NHS Foundation Trust and the University of Oxford. AR is funded by a fellowship from MQ: Transforming Mental Health.

## Notes

### Competing Interest Statement

The authors have declared no competing interest.

### Funding Statement

This study was funded by the JABBS Foundation

### Author Declarations

Written informed consent was obtained from all participants prior to the start of the study. Study approval was granted by South Central Oxford REC B (Ref: 118784, Ethics number: 12/SC/0713).

