## Supplementary materials for "Brain activity measured by functional brain imaging predicts breathlessness improvement during pulmonary rehabilitation"

Michael Browning

Eugene Duff

Catherine J. Harmer

Andrea Reinecke

Najib M. Rahman

Kyle T.S. Pattinson

**Online Data Supplement**

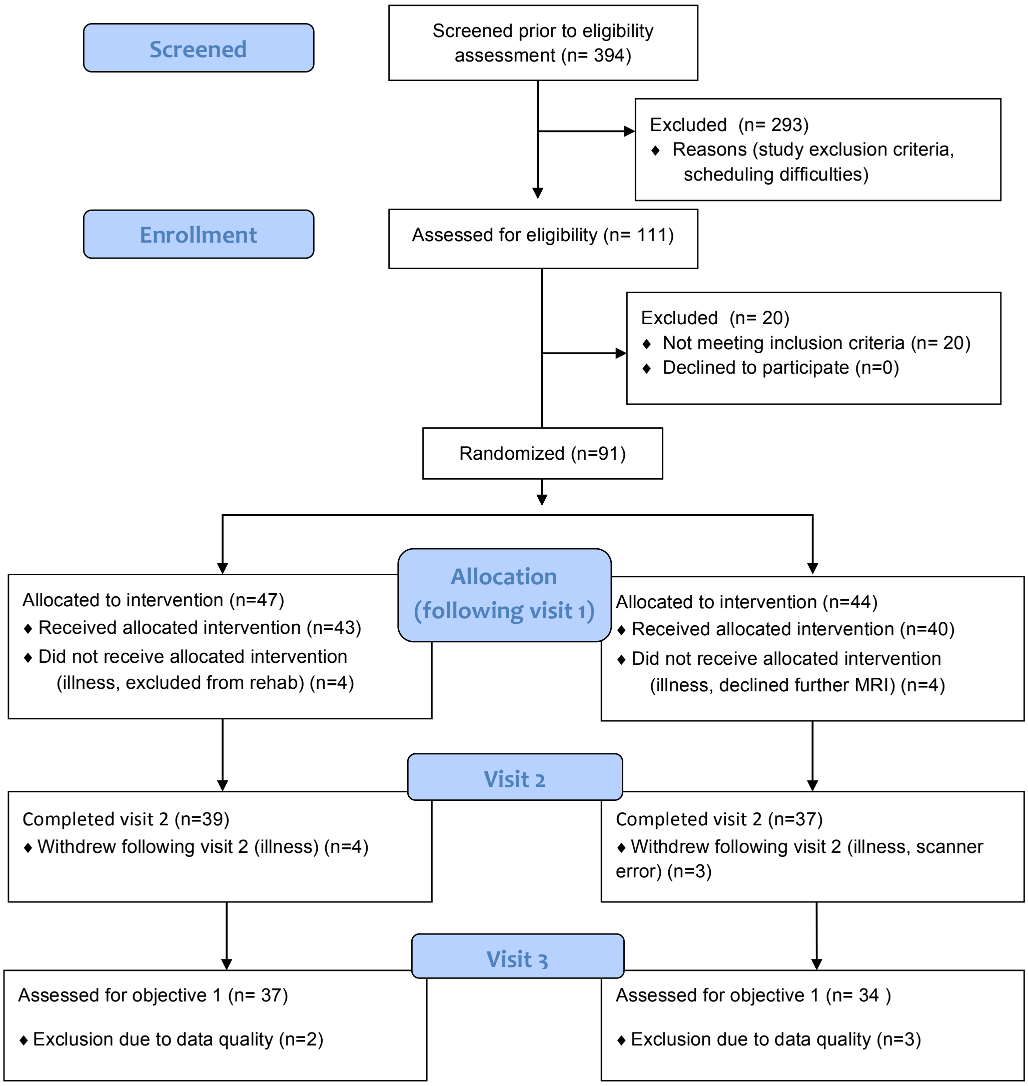


***Supplementary Figure 1.*** *Consort diagram illustrating stages of participant recruitment from initial screening through to completion at visit 3.*

91 participants (30 female, median age 70 years; range 46-85 years) with COPD were recruited to this study. 72 participants completed all three visits. 1 participant's brain imaging data was lost due to data collection error.

#### *Inclusion criteria*

Study inclusion criteria were: a diagnosis of COPD and admittance to pulmonary rehabilitation. Exclusion criteria were: inadequate understanding of verbal and written

English, significant cardiac, psychiatric (including depression under tertiary care) or metabolic disease (including insulin-controlled diabetes), stroke, contraindications to either D-cycloserine (including alcoholism) or magnetic resonance imaging (MRI), epilepsy, claustrophobia, regular therapy with opioid analgesics or home oxygen therapy.

***Table 1.*** *Demographic information from the 72 participants who completed all study visits. Variance is expression either in terms of standard deviation (SD) or interquartile range (IQR) depending on the normality of the underlying data distribution.* ***BMI*** *= Body Mass Index,* ***MRC*** *= Medical Research Council.* ***$SpO_2$%*** *= Peripheral Oxygen saturation, expressed as a percentage. Also listed with prevalence in brackets are recorded comorbidities ordered by frequency*

| | Pre-rehabilitation | Post-rehabilitation |
|---|---|---|
| Age (median years / range) | **71 / (46-85)** | |
| Smoking pack-years (years / IQR) | **30.1 ± 28.5** | |
| BMI (kg.m-2 ± SD) | **26.9 ± 6.0** | **27.4 ± 4.6** |
| MRC breathlessness scale (IQR) | **3 (1)** | **2 (1)** |
| Resting $SpO_2$% (IQR) | **95 ± 3.75** | **94 ± 3.75** |
| Resting heart rate (beats.min-1 ± SD) | **80.8 ± 14.1** | **78.3 ± 13.0** |
| FEV1/FVC (IQR) | **0.55 (0.15)** | **0.57 (0.28)** |
| Duration of breathlessness (years / IQR) | **10 (15.6)** | |
| Total exacerbations (number / IQR) | **0 (2.3)** | |

**Comorbidities (frequency)**

| | | |
|---|---|---|
| Asthma (25) | Reflux and heart burn (22) | Hypertension (24) |
| Swelling of both ankles (19) | Surgery to the chest (13) | Depression (13) |
| Diabetes (9) | Heart attack (9) | Bronchiectasis (7) |
| Osteoporosis (6) | Arrhythmia (6) | Inflammatory bowel disease (5) |
| Peptic ulcer (5) | Heart failure (2) | Neuromuscular weakness (2) |
| Tuberculosis (1) | | |

***Table 2.*** *Demographic information from the 72 participants who completed the three study visits expressed as the group total group and D-cycloserine and placebo groups. Variance is expression either in terms of standard deviation (SD) or interquartile range (IQR).* ***BMI*** *= Body Mass Index,* ***MRC*** *= Medical Research Council clinical measure of breathlessness.* ***$SpO_2$%*** *= Peripheral Oxygen saturation, expressed as a percentage.*

| **Visit 1 (N=72)** | Total | D-cycloserine | Placebo |
|---|---|---|---|
| Age (median years/range) | **71 / (46-85)** | **71 / (47-81)** | **71.5 / (46-85)** |
| Smoking pack-years (IQR) | **30 / (28.5)** | **34 / (25.6)** | **30 / (30.0)** |
| BMI kg.m-2 ± SD | **26.9 ± 6.0** | **27.3 ± 6.5** | **26.9 ± 5.7** |
| MRC (IQR) | **3 (1)** | **3 (1)** | **2.5 (1)** |
| Resting $SpO_2$% (IQR) | **95 / (3.8)** | **95 / (3.3)** | **94.5 / (3.0)** |
| Resting heart rate beats.min-1 ± SD | **80.8 ± 14.1** | **80.8 ± 13.4** | **80.8 ± 15.0** |
| FEV1/FVC (IQR) | **0.55 / (0.15)** | **0.53 / (0.17)** | **0.56 / (0.13)** |

**Study Drug**

Study drugs were purchased from Ipswich Hospital Pharmacy Manufacturing Unit, Heath Road, Ipswich IP4 5PD, Tel: 01473 703603.

**Randomisation Procedure**

Once the participant gave written consent to the trial and completed the MRI scan, a member of the team submitted a randomisation form, entering eligibility criteria and minimisation factors. Allocation to active or placebo capsules was carried out by Sealed Envelope Randomisation Services (Sealed Envelope Ltd, Concorde House, Grenville Place, London NW7 3SA). The randomisation number was then provided to the Oxford Respiratory Trials Unit who dispensed the drug/placebo. Minimisation factors were as follows:

1. Centre
2. MRC grade
3. Diabetes
4. Antidepressant
5. Age at which the participant completed full time education
6. Previous rehabilitation

Randomisation codes were held by Sealed Envelope until study completion, after which at the first stage of unblinding an independent researcher provided study researchers with a coded binarised system for analysis. Researchers remained blinded to group identity until analysis was completed. No side effects were reported.

***Study Visit Protocol***

Following telephone screening participants were invited to attend their first research session (baseline) prior to starting pulmonary rehabilitation. Pulmonary rehabilitation courses were run by either Oxford Health NHS Foundation Trust, West Berkshire NHS Foundation Trust, or Milton Keynes University Hospitals NHS Trust. Following the successful completion of the first study session participants were then randomised to receive either the study drug or placebo. Participants then attended their first four sessions of pulmonary rehabilitation, 30 minutes prior to each of these sessions they received their assigned study drug or placebo tablet. A second study visit took place following the fourth pulmonary rehabilitation session but before the 6th session. Participants then completed the remainder of their pulmonary rehabilitation course before attending a third study session (Supplementary Figure 2) that occurred in the two weeks after termination of pulmonary rehabilitation. For the purposes of this study, data were used from visits occurring before (visit one) and following the completion of pulmonary rehabilitation (visit three).

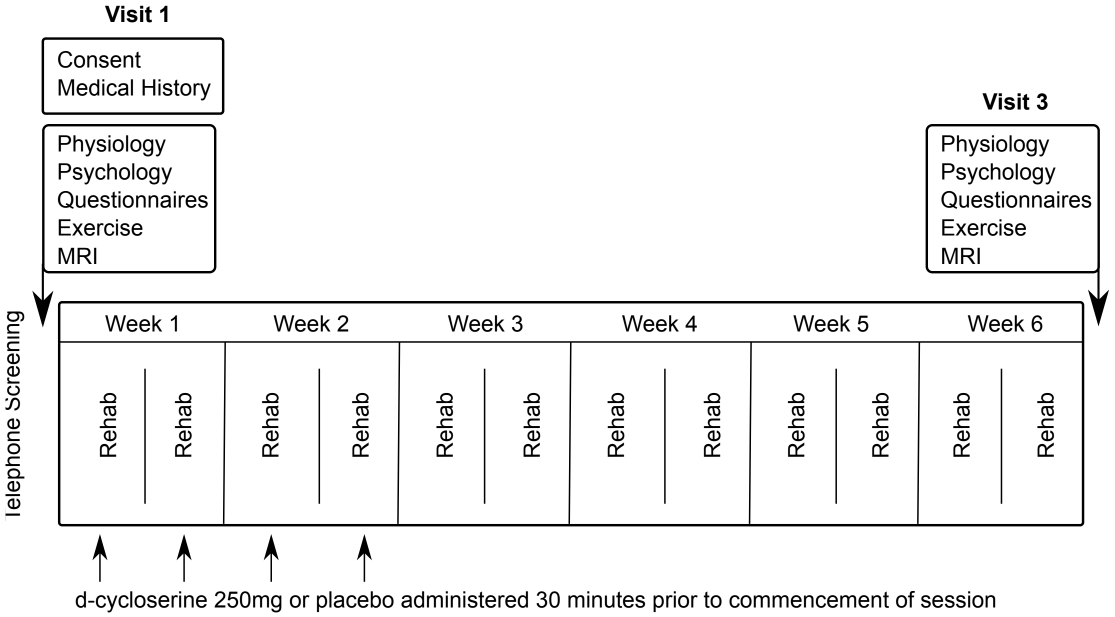


***Supplementary Figure 2.*** *A schematic demonstrating order of visits, rehabilitation sessions and tablet administration throughout the study period. Participants took part in one study visit prior to their first pulmonary rehabilitation session. Study drug/placebo were administered on four occasions over the first four rehabilitation sessions. Participants continued with their pulmonary rehabilitation course for a further four weeks before returning for a final visit.*

### Behavioural Measures

*Questionnaire Measures*

**Dyspnoea-12 (D12) Questionnaire**: This is a 12-item questionnaire designed to measure the severity of breathlessness and has been validated for use in patients with respiratory disease [1].

**Centre for Epidemiologic Studies Depression Scale (CES-D)**: Depressive symptoms are commonly observed in patients with respiratory disease. This brief questionnaire consists of 20 items investigates the symptoms of depression across a number of factors [2].

**State-Trait Anxiety Inventory (STAIT-T)**: This questionnaire assesses participant's general level of anxiety in particular scenarios via 20 questions asking "how anxious you generally feel" [3].

**Fatigue Severity Scale**: This 9-point questionnaire quantifies patient fatigue, which is well documented in its association with COPD [4].

**St George's Respiratory Questionnaire (SGRQ)**: There are 50 questions in this questionnaire, which has been developed and validated for use in COPD and asthma. The questions measure the impact of overall health, daily life and well-being [5].

**Medical Research Council (MRC) breathlessness scale**: The MRC scale quantifies perceived difficulty due to respiratory restrictions on a scale of 1 to 5 [6].

**Mobility Inventory (MI)**: This questionnaire collects data regarding the extent to which a participant avoids certain situations, either alone or accompanied (21-items in each category) [7].

**Breathlessness Catastrophising Scale:** Adapted from Catastrophic Thinking Scale in Asthma. This 13-point questionnaire was modified for this study by substituting the word "asthma" for "breathlessness" in order to measure catastrophic thinking [8] [9].

**Breathlessness Awareness and Vigilance Scale Pain:** Adapted from Pain Awareness and Vigilance Scale. This questionnaire was modified by substituting the word "breathlessness" for the word "pain". The 16-point scale measures how much a participant focuses their attention onto their breathlessness [10] [9].

**Physiological Measures**

A trained respiratory nurse collected spirometry measures of $FEV_1$ and FVC using Association for Respiratory Technology and Physiology standards [11]. Participants performed two modified incremental shuttle walk tests (MSWT) [12], and heart rate and oxygen saturations ($SpO_2$) were measured immediately before the MSWT and subsequently every minute until 10 minutes post-exercise (or until participants returned to their baseline state) using a fingertip pulse oximeter ($Go_2$; Nonin Medical Inc). Before and after the MWST participants also rated their breathlessness on a modified Borg scale [13]. In a MWST participants must walk between and around two cones, placed 10m apart in time to a set of auditory beeps played from a laptop. Initially the speed of beep repetition is slow, but the participant must increase their walking speed each minute in order to reach the cone before the next beep. Participants continue to walk (or run) until they are too breathless to continue, at which point the total distance walked is recorded.

**MRI Acquisition**

Prior to each MRI session participants were screened for standard MRI contraindications including metal in or about their person, epilepsy and claustrophobia.

*Image acquisition:*

Hardware: A Tim System (Siemens Healthcare GmbH) 12-channel head coil.

T1 sequence parameters: TR, 2040ms; TE, 4.68ms; voxel size, 1 x 1 x 1 mm; FOV, 200mm; flip angle, 8°; inversion time, 900ms; bandwidth 130 Hz/Px).

T2*-weighted (functional) sequence parameters: TR, 3000ms; TE 30ms; voxel size 3 x 3 x 3 mm; FOV, 192mm; flip angle 87°; echo spacing 0.49ms.

Functional scan durations: word-task - 215 volumes, 7 minutes and 33 seconds duration.

Field map scans of the $B_0$ field were obtained to aid the distortion correction of the functional scans: TR, 488ms; TE1, 5.19ms; TE2, 7.65ms; flip angle 60°; voxel size, 3.5 x 3.5 x 3.5 mm.

*Word Task*

This task was developed and published by Herigstad and colleagues in 2016 for use in the COPD population [14]. Word cues were developed in three key stages; firstly in collaboration with respiratory practitioners, academics and physiotherapists, a set of 30 word cues associated with breathlessness were created. Next, these cues were provided to patients with COPD alongside a VAS rating scale, allowing patients to rate how breathless and anxious the situations identified by the cues would make them feel. Following adjustments based on participant feedback, the word cues were then computerised and tested in a larger population of COPD patients [14]. Further validation was carried out in the fMRI environment and by for clinical sensitivity with comparisons between changes in key questionnaire measures and word-cue rating. Before the first scan session, participants were given the opportunity to practice using the button box with a set of test words.

In this task brain activity was correlated with corresponding visual analogue ratings of anxiety and breathlessness. During the fMRI scanning, participants were presented with a word cue in white text on a black background for 7 seconds. Participants were then asked, “how breathless would this make you feel” (wB) and “how anxious would this make you feel” (wA). To each question participants responded within a 7 second window using a button box and visual analogue scale (VAS). The response marker always initially appeared at the centre of the scale, with the anchors “Not at all” and “Very much” at either end. A control condition, used as a baseline measure of activity in response to the presentation of a visual stimulus was presented 4 times over the course of the scan, consisting of a string of “XXXXXXXXXXXXXXX” with fixed length of 15 characters, and each time was presented for 7 seconds. No rating period followed these control blocks [14].

#### ***Imaging Analysis***

*Functional MRI Preprocessing*

Data denoising was carried out as follows: Before the first level analysis, each functional scan was decomposed into maximally independent components using

FMRIB's MELODIC tool (Multivariate Exploratory Linear Optimised Decomposition into Independent Components). "Noise" components were identified by FIX (FMRIB's auto-classification tool, [15, 16]) using the WhII.Standard.RData [17] trained classifier with aggressive clean up option. A Principle Component Analysis (PCA) was run on the FIX identified components to retrain 99% of the variance. Separately, the cardiac and respiratory related physiological signals (recorded via a pulse oximeter and a respiratory bellows) were transformed into a series of regressors, (three cardiac and four respiratory harmonics) as well as an interaction term and a measure of respiratory volume per unit of time (RVT), using FSL's physiological noise modelling tool (PNM). The signal associated with these waveforms (modelled using retrospective image correction (RETROICOR) [18, 19]) was then used to form voxelwise noise regressors.

The confounds identified by FSL's FIX and PNM tools, along with sources of noise arising from motion, were then combined into a single model. This single noise model approach builds upon the technique outlined by [20]; and fully detailed by [21]. In these preceding works we employed a step-wise technique whereby physiological noise (identified by PNM) and FIX-identified noise were each removed from the data in separate steps prior to data entry into the lower level model. In the new cleanup pipeline, a single text file containing time-course information relating to FIX identified noise components along with white matter or CSF related noise was included as additional confound EV's within the lower level model, while the PNM-identified noise was entered into the model as a standard voxel-wise confound list. In this updated de-noising pipeline, confounds identified above are added to model at the stage of first-level analysis and thus the functional dataset can be corrected for sources of noise arising from motion, scanner and cerebro-spinal fluid artefacts, cardiac, and respiratory noise in a single step, rather than three.

*Functional MRI Analysis*

MRI processing was performed using FEAT (FMRI Expert Analysis Tool within the FSL package). The data were corrected for movement using MCFLIRT (Motion correction using FMRIB's Linear Image Registration Tool [22]). Non-brain structures were removed using BET (Brain Extraction Tool [23]). Spatial smoothing was carried out using a full-width-half-maximum Gaussian kernel of 5mm, while high-pass temporal filtering (Gaussian-weighted least squares straight line fitting; 90 s) removed low frequency noise and slow-drift. Distortion correct of EPI data was carried out using a combination of FUGUE (FMRIB's Utility for Geometrically Unwarping EPI's [24, 25] and BBR (Boundary Based Registration; part of the FMR Expert Analysis Tool, FEAT version 6.0 [26]). The data were corrected for physiological noise using FSL's FIX-PNM pipeline. Functional scans were registered in a two-step process to the MNI152 (1x1x1 mm) standard space brain template. Firstly, each subject's EPI was registered to their associated T1-weighted structural image using BBR (6 DOF) with nonlinear field map distortion correction [26]. In the second step the subject's structural image was registered to 1mm standard space via an affine transformation followed by nonlinear registration (using FNIRT: FMRIB's Non-linear Registration Tool [27]).

***First Level Processing***

*Functional MRI: Word-cue task*: At the individual subject level, a general linear model (GLM) was created with explanatory variables (EVs) for breathlessness word or non-word presentation, and two de-meaned EVs modeling the reported breathlessness and anxiety response to the word cues (Supplementary Figure 3). Additional explanatory noise variables were included to model the period during which the participant responded using the visual analog scale (VAS).

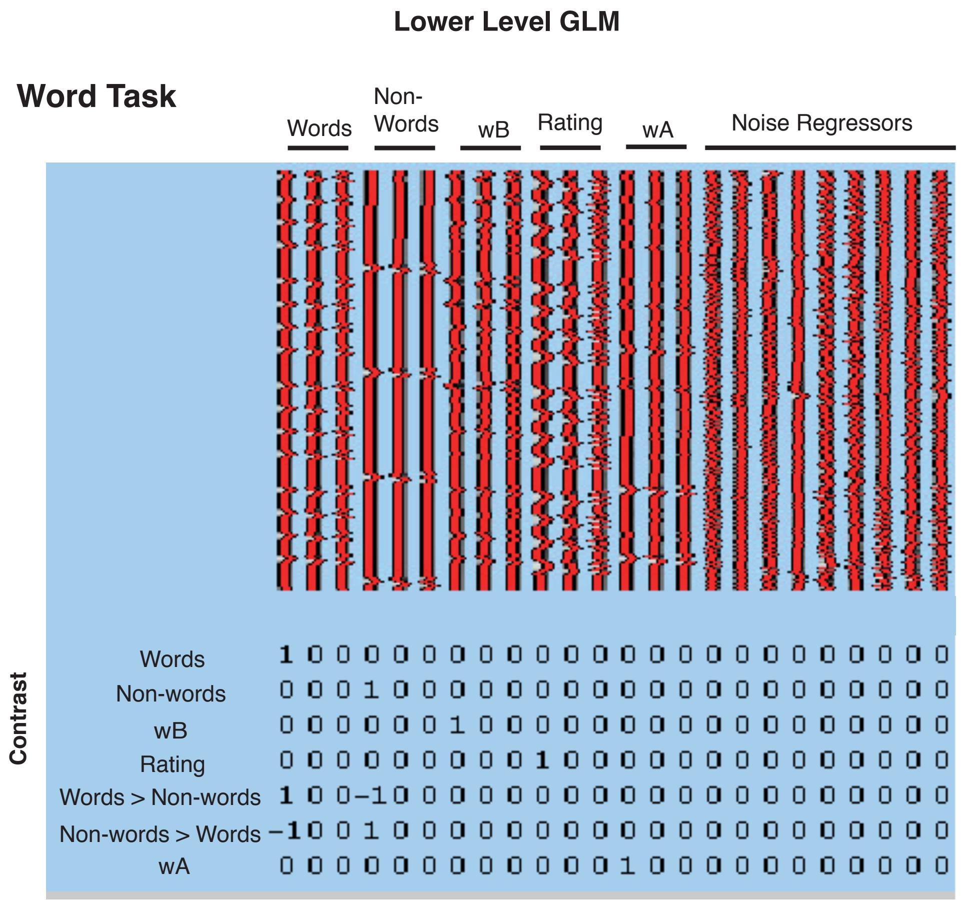


***Supplementary Figure 3*** *– An illustration of the generalised linear models (GLM) used lower level analyses for the word task. Abbreviations as follows – wB – breathlessness rating, wA – breathlessness anxiety rating.*

### Regions of Interest

Regions of interest defined by the Harvard-Oxford Atlas and Destrieux' cortical atlas are listed here with atlas label identifier in brackets if different from anatomical name.

1. Anterior Insular Cortex (G_insular_short)
2. Posterior Insular Cortex (G_Ins_lg_and_S_cent_ins)
3. Anterior Cingulate Cortex (Cingulate Cortex, anterior division)
4. Amygdala
5. Hippocampus
6. Posterior Cingulate Cortex (Cingulate Cortex, posterior division)

7. Medial Prefrontal Cortex (Frontal Pole)
8. Middle frontal Gyrus
9. Superior Marginal Gyrus (Supramarginal Gyrus)
10. Superior Frontal Gyrus
11. Putamen
12. Precuneus
13. Angular Gyrus
14. Caudate
15. Precentral Gyrus

A 40% probability threshold was applied to the mask of each region. The regions were then registered to each individual before being re-thresholded at 40% probability to avoid interpolation errors and were binarized.

**Model specifics and technical definitions**

**Cross validation –** Is a resampling procedure. Data can be split into a number of different training and testing folds (k-folds). This enables the algorithm to learn from the maximum number of new data points.

**Support Vector Machines (SVM) –** Separates pre-defined classes by establishing an optimal boundary within high-dimensional space called a hyper-plane. In this work we used a linear kernel which draws an assumption on the relationship between activity space and feature space but places no assumption on the distribution that the two are drawn from.

**Elastic net regularisation –** Model fitting involves a trade-off between bias and variance. Bias is the difference between predicted regression parameters and actual estimator, essentially a measure of accuracy. Variance is the uncertainty of those estimations. Both of these numbers should be low. Regularisation provides a way to reduce the variance while introducing some bias. Elastic net regularisation combines elements of ridge and lasso regression techniques. Ridge elements - A cost function is applied to the slope of any fit function to reduce sudden changes, helping with the stability of the classifier. Lasso elements – accounts for magnitude of feature contribution to the classifier but struggles where variables are highly correlated. Elastic net includes terms for both slope and magnitude as a mix-ratio which encourages groupings of variables.

**Random OverSample Examples (ROSE) –** Imbalanced classes can affect classifier performance. ROSE creates an artificially balanced sample using a smoothed bootstrap approach.

**Seed setting –** Random seeds are used in the generation of models. The random number is set locally each time to the same number to ensure the model sequence is reproducible.

**Confusion matrices –** A visual representation of the success of a supervised classifier. Where rows represent the classifiers attempt and columns represent the actual class.

| | | Actual Class | |
|---|---|---|---|
| Predicted Class | | Dog | Cat |
| | Dog | **5** | 2 |
| | Cat | 3 | **3** |

### Results

A

B

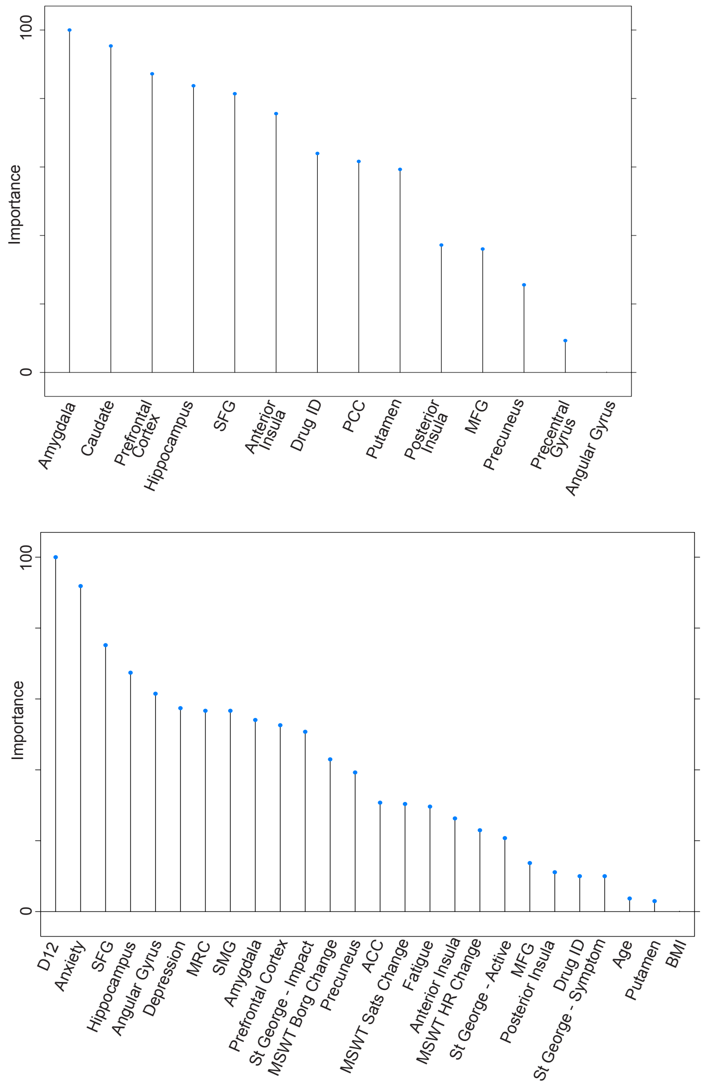


***Supplementary Figure 4.*** *Needle plot of ranked importance for each of the* ***(A)*** *brain derived metrics (full model) and* ***(B)*** *brain derived metrics and non-imaging measures (full model), to the classification of responder/non-responders. Abbreviations: SFG – Superior Frontal Gyrus, PCC – Posterior Cingulate Cortex, MFG – Middle Frontal Gyrus, ACC – Anterior Cingulate Cortex, MSWT – Modified Shuttle Walk Test, HR – Heart Rate, BORG – breathlessness scale, BMI – Body Mass Index, Sats – Oxygen saturation.*

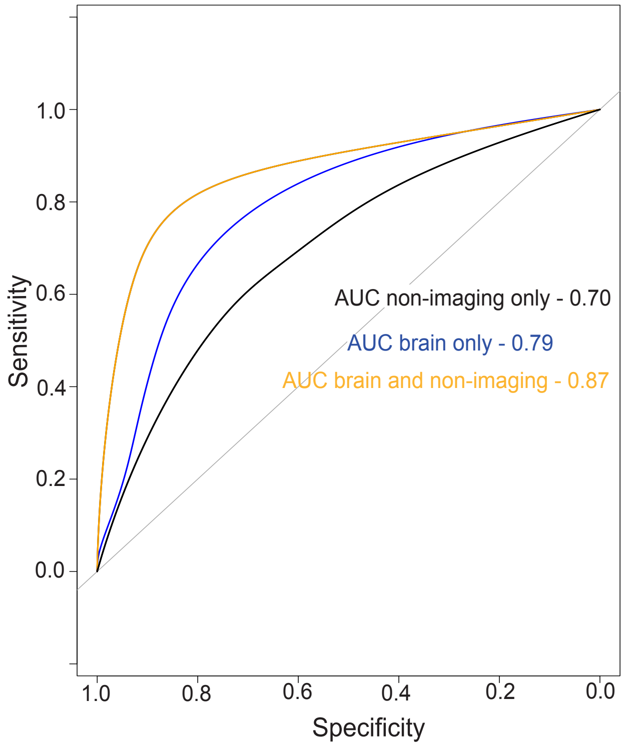


***Supplementary Figure 5.*** *Receiver Operator Curves for each of the three full models. AUC – area under the curve.*


1. Yorke, J., et al., *Quantification of dyspnoea using descriptors: development and initial testing of the Dyspnoea-12.* 2010. **65**(1): p. 21-26.
2. Radloff, L.S., *The CES-D Scale:A Self-Report Depression Scale for Research in the General Population.* 1977. **1**(3): p. 385-401.

3. Spielberger, C.D., *State-Trait Anxiety Inventory*, in *The Corsini Encyclopedia of Psychology*. 2010.
4. Krupp, L.B., et al., *The fatigue severity scale: Application to patients with multiple sclerosis and systemic lupus erythematosus.* Archives of Neurology, 1989. **46**(10): p. 1121-1123.
5. Jones, P.W., et al., *A self-complete measure of health status for chronic airflow limitation. The St. George's Respiratory Questionnaire.* Am Rev Respir Dis, 1992. **145**(6): p. 1321-7.
6. Bestall, J.C., et al., *Usefulness of the Medical Research Council (MRC) dyspnoea scale as a measure of disability in patients with chronic obstructive pulmonary disease.* Thorax, 1999. **54**(7): p. 581-586.
7. Chambless, D.L., et al., *The Mobility Inventory for Agoraphobia.* Behav Res Ther, 1985. **23**(1): p. 35-44.
8. De Peuter, S., et al., *Illness-specific catastrophic thinking and overperception in asthma.* Health Psychol, 2008. **27**(1): p. 93-9.
9. Herigstad, M., et al., *Dyspnea-related cues engage the prefrontal cortex: evidence from functional brain imaging in COPD.* Chest, 2015. **148**(4): p. 953-961.
10. McCracken, L.M., K.E. Vowles, and C. Eccleston, *Acceptance-based treatment for persons with complex, long standing chronic pain: a preliminary analysis of treatment outcome in comparison to a waiting phase.* Behav Res Ther, 2005. **43**(10): p. 1335-46.
11. Levy, M.L., et al., *Diagnostic spirometry in primary care: Proposed standards for general practice compliant with American Thoracic Society and European Respiratory Society recommendations: a General Practice Airways Group (GPIAG)1 document, in association with the Association for Respiratory Technology & Physiology (ARTP)2 and Education for Health3 1 www.gpiag.org 2 www.artp.org 3 www.educationforhealth.org.uk.* Prim Care Respir J, 2009. **18**(3): p. 130-47.
12. Bradley, J., et al., *Validity of a modified shuttle test in adult cystic fibrosis.* Thorax, 1999. **54**(5): p. 437-9.
13. Mahler, D.A., et al., *Comparison of clinical dyspnea ratings and psychophysical measurements of respiratory sensation in obstructive airway disease.* Am Rev Respir Dis, 1987. **135**(6): p. 1229-33.
14. Herigstad, M., et al., *Development of a dyspnoea word cue set for studies of emotional processing in COPD.* Respiratory physiology & neurobiology, 2016. **223**: p. 37-42.
15. Griffanti, L., et al., *ICA-based artefact removal and accelerated fMRI acquisition for improved resting state network imaging.* Neuroimage, 2014. **95**: p. 232-47.
16. Salimi-Khorshidi, G., et al., *Automatic denoising of functional MRI data: combining independent component analysis and hierarchical fusion of classifiers.* Neuroimage, 2014. **90**: p. 449-68.
17. Filippini, N., et al., *Study protocol: the Whitehall II imaging sub-study.* BMC Psychiatry, 2014. **14**(1): p. 159.
18. Harvey, A.K., et al., *Brainstem functional magnetic resonance imaging: disentangling signal from physiological noise.* J Magn Reson Imaging, 2008. **28**(6): p. 1337-44.

19. Brooks, J.C., et al., *Physiological noise modelling for spinal functional magnetic resonance imaging studies.* Neuroimage, 2008. **39**(2): p. 680-92.
20. Faull, O.K., et al., *Conditioned respiratory threat in the subdivisions of the human periaqueductal gray.* Elife, 2016. **5**.
21. Hayen, A., et al., *Opioid suppression of conditioned anticipatory brain responses to breathlessness.* Neuroimage, 2017. **150**: p. 383-394.
22. Jenkinson, M., et al., *Improved optimization for the robust and accurate linear registration and motion correction of brain images.* Neuroimage, 2002. **17**(2): p. 825-41.
23. Smith, S.M., *Fast robust automated brain extraction.* Hum Brain Mapp, 2002. **17**(3): p. 143-55.
24. Holland, D., J.M. Kuperman, and A.M. Dale, *Efficient correction of inhomogeneous static magnetic field-induced distortion in Echo Planar Imaging.* Neuroimage, 2010. **50**(1): p. 175-83.
25. Jenkinson, M., *Fast, automated, N-dimensional phase-unwrapping algorithm.* Magn Reson Med, 2003. **49**(1): p. 193-7.
26. Greve, D.N. and B. Fischl, *Accurate and robust brain image alignment using boundary-based registration.* Neuroimage, 2009. **48**(1): p. 63-72.
27. Andersson, J., *Non-Linear registration, aka spatial normalisation.* FMRIB technical report, 2010. **TR07JA2**.
